# Precision Breast Cancer Screening with a Polygenic Risk Score

**DOI:** 10.1101/2020.08.17.20176263

**Authors:** Tõnis Tasa, Mikk Puustusmaa, Neeme Tõnisson, Berit Kolk, Peeter Padrik

## Abstract

Breast cancer (BC) is the leading cause of cancer deaths in women in the world. Genome-wide association studies have identified numerous genetic variants (SNPs) independently associated with BC. The effects of such SNPs can be combined into a single polygenic risk score (PRS). Stratification of women according to PRS could be introduced to primary and secondary prevention. Our aim was to revalidate a PRS model and to develop a pipeline for individualizing breast cancer screening.

Previously published PRS models for predicting the risk of breast cancer were collected from the literature. Models were validated on the Estonian Biobank (EGC) dataset consisting of 32,548 quality-controlled genotypes with 315 prevalent and 365 incident BC cases and on 249,062 samples in the UK Biobank dataset consisting of 8637 prevalent and 6825 incident cases. The best performing model was selected based on the AUC in prevalent data and independently validated in both incident datasets. Using Estonian BC background information, we performed absolute risk simulations and developed individual risk-based recommendations for prevention.

The best-performing PRS included 2803 SNPs. The C-index of the Cox regression model associating BC status with PRS was 0.656 (SE = 0.05) with a hazard ratio of 1.66 (95% confidence interval 1.5 - 1.84) on the incident EGC dataset. The PRS is able to stratify individuals with more than a 3-fold risk increase. The observed 10-year risks of individuals in the 99th percentile exceeded the 1^st^ percentile more than 10-fold.

PRS is a powerful predictor of breast cancer risk. Currently, PRS scores are not implemented in routine BC screening. We have developed PRS-based recommendations for personalized primary and secondary prevention and our approach is easily adaptable to other nationalities by using population-specific background data of other genetically similar populations.

## Introduction

Breast cancer (BC) is the leading cause of cancer deaths in women. Every year adds 2 million new diagnoses and more than 600 000 deaths (1). Among women of European ancestry, the probability of BC onset before the age of 85 is approximately 1 in 8, and 20% of cases occur in women younger than 50 (2). Around 30% of the total BC risk has been shown as hereditary (3). Genetic factors include pathogenic mutations in high and moderate-risk cancer predisposition genes (*BRCA1, BRCA2, ATM, CHEK2, TP53, PTEN, STK11, CDH1, PALB2, NBN, NF1*, and *BARD1*), having effects large enough to warrant monogenic testing (4–6). However, only a fraction (5-10%) of BC cases are caused by these rare genetic variants (7). A considerable part of BC variation is explained by variants outside these high-risk genes in the form of BC-associated common single-nucleotide polymorphisms (SNPs) (8). It has only recently become possible to aggregate information across many common SNPs to predict disease risk and to develop applications with potential clinical utility.

Several studies have combined SNPs with genome-wide significance into a summary estimate of BC risk using a polygenic risk score (PRS) approach (9–11). Initial attempts by Mavaddat *et al*. (10) and Sieh *et al*. (12) demonstrated a strong effect of the score in predicting future BC cases. Other major efforts such as those by Mavaddat *et al*., Khera *et al*. and Hughes *et al*. additionally elucidated the use of PRS as a BC predictor (9, 13, 14).

BC screening with mammography reduces breast cancer mortality risk 20-40% (15–17). Current BC screening guidelines in Estonia are based on age and do not support regular screening of women below the age of 50. This protocol disregards younger women with a higher genetic risk. Breast cancer PRSs identify differences in genetic risks and provide a straightforward basis for designing personalized screening programs by accounting for individual genetic susceptibility (18). Currently, PRS scores have not been implemented in routine BC screening but simulations have suggested that risk profile informed behavioral adjustments could provide cost-savings to screening and guide risk-based followup actions (19, 20). High-risk estimation could be also the indication for the use of hormonal chemoprevention (21).

This study aims to evaluate the risk prediction performance of several published PRS and to assess its use as a risk stratification approach in the context of Estonia. Concretely, we aim to use information from polygenic risk stratification to propose individual follow-up actions based on current screening onboarding strategies.

## Methods

### Biobank participant data

BC datasets were acquired from two population biobanks: the Estonian Biobank of the Estonian Genome Center at the University of Tartu (EGC) and the UK Biobank (UKBB). Quality controlled samples were divided into prevalent and incident datasets. The prevalent dataset included BC cases diagnosed before Biobank recruitment with 5 times as many controls without the diagnosis. Incident data included cases diagnosed in any of the linked databases after recruitment to the Biobank and all controls not included in the prevalent dataset. Prevalent datasets were used for identifying the best candidate model and the incident datasets were used to obtain an independent PRS effect estimate on BC status.

### Participant data of Estonian Genome Center

BC cases and controls in retrospective data of EGC were defined by breast cancer ICD-10 code (C50) status derived from questionnaires filled at recruitment of the gene donors and from linked data from Estonian Cancer Registry (data until 2013), National Health Insurance Fund (data until the end of 2018) and Causes of Death Registry (data until 2017 May).

All EGC samples were genotyped in Core Genotyping Lab of Institute of Genomics, University of Tartu, using Illumina GSAMD-24v1-0 arrays. Individuals were excluded if their call-rate was < 95% or sex defined based on X chromosome heterozygosity did not match declared sex. Variants were filtered by call-rate < 95% on the whole EGC dataset, Hardy-Weinberg equilibrium (HWE) test p-value < 1e-4 (autosomal variants only) and minor allele frequency < 1%. Variant positions were updated to b37 and all variants were changed to the TOP strand (https://www.well.ox.ac.uk/~wrayner/strand/). Phasing was done using Eagle (v. 2.3) software (22) and imputation with Beagle (v. 28Sep18.793) (23) using the Population-specific imputation reference of 2297 WGS samples (24).

### Participant data of UK Biobank

This study used genotypes from the UK Biobank cohort (version v3, obtained 07.11.2019) and made available to Antegenes under application reference number 53602. The data was collected, genotyped using either the UK BiLEVE or Affymetrix UK Biobank Axiom Array. Breast cancer cases in the UK Biobank cohort were retrieved by the status of ICD-10 code C50. We additionally included cases with self-reported UK Biobank code “1022”.

Quality control steps and in detail methods applied in imputation data preparation have been described by the UKBB and made available at http://www.ukbiobank.ac.uk/wpcontent/uploads/2014/04/UKBiobank_genotyping_QC_documentation-web.pdf. We applied additional quality controls on autosomal chromosomes. First, we removed all variants with allele frequencies outside 0.1% and 99.9%, genotyping call rate < 0.1, imputation (INFO) score < 0.4 and Hardy-Weinberg equilibrium test p-value < 1E-6. Sample quality control filters were based on several pre-defined UK Biobank filters. We removed samples with excessive heterozygosity, individuals with sex chromosome aneuploidy, and excess relatives (> 10). Additionally, we only kept individuals for whom the submitted gender matched the inferred gender, and the genotyping missingness rate was below 5%.

### Model selection from candidate risk models

We searched the literature for PRS models in the public domain. The requirements for inclusion to the candidate set were the availability of the chromosomal location, reference and alternative allele, minor allele frequency, and an estimator for the effect size either as odds ratio (OR) or its logarithm (log-OR) specified for each genetic variant. In cases of iterative model developments on the same underlying base data, we retained chronologically newer ones. The search was performed with Google Scholar and PubMed web search engines by working through a list of articles using the search [“Polygenic risk score” or “genetic risk score” and “breast cancer”], and then manually checking the results for the inclusion criteria. We additionally pruned the PRS from multi-allelic, non-autosomal, non-retrievable variants based on bioinformatics re-analysis with Illumina GSA-24v1 genotypes and non-overlapping variants between EGC and UKBB data.

PRSs were calculated as 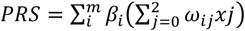, where ω_ij_ is the probability of observing genotype *j*, where *j* ∈ {0, 1, 2} for the i-th SNP; *m* is the number of SNPs, and *β_i_* is the effect size of the i-th SNP estimated in the PRS. The mean and standard deviation of PRS in the cohort were extracted to standardize individual risk scores to Gaussian. We tested the assumption of normality with the mean of 1000 Shapiro-Wilks test replications on a random subsample of 1000 standardized PRS values.

Next, we evaluated the relationship between BC status and standardized PRS in the two prevalent datasets with a logistic regression model to estimate the logistic regression-based odds ratio per 1 standard deviation of PRS (*OR_sd_*), its p-value, model Akaike information criterion (AIC) and Area Under the ROC Curve (AUC). The logistic regression model was compared to the null model using the likelihood ratio test and to estimate the Nagelkerke and McFadden pseudo-R^2^. We selected the candidate model with the highest AUC to independently assess risk stratification in the incident datasets.

### Independent performance evaluation of a polygenic risk score model

The main aim of the analyses in the incident datasets was to derive a primary risk stratification estimate, hazard ratio per 1 unit of standardized PRS (HR*sd*), using a right-censored and left-truncated Cox-regression survival model. The start of time interval was defined as the age of recruitment; follow-up time was set as the time of diagnosis for cases and at the time of last health data linkage for controls. Scaled PRS was used as the only independent variable of BC diagnosis status. 95% confidence intervals were created using the standard error of the log-hazard ratio. We also assessed the goodness-of-fit of the survival model using Harrell’s C-index and the likelihood ratio test.

Further, we evaluated the concordance between theoretical hazard ratio estimates derived with the continuous per unit PRS (HRsd) estimate and the hazard ratio estimates inferred empirically from data. For this, we binned the individuals by PRS to 5%-percentiles and estimated empiric hazard ratio of BC directly between those classified in each bin and those within the 40-60 PRS percentile. Theoretically estimated hazard ratio estimates assume a multiplicative effect of the mean in a PRS bin on the unit based hazard ratio. This relationship between HRsd and the expected mean in the truncated Gaussian PRS distribution is expressed as 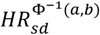, where the exponent is the mean of a truncated Gaussian distribution between two percentiles *a* and *b* (bounded between 0 and 1, a<b), and 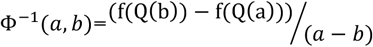, where *Q*(*b*) is the Gaussian quantile function on a percentile *b* and *f*(*Q*(*b*)) is the Gaussian probability density function value at a quantile function value. We compared the two approaches by using the Spearman correlation coefficient and the proportion of distribution-based 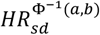 estimates in empirical confidence intervals.

### Absolute risk estimation

Individual τ-year (e.g. 10-year) absolute risk calculations are based on the risk model developed by Pal Choudhury et al. (25). Individual absolute risks are estimated for currently a-year old individuals in the presence of known risk factors (*Z*) and their relative log hazard-ratio parameters (*β*). 95% uncertainty intervals for the hazard ratio were derived using the standard error and z-statistic 95% quantiles CIHR = exp(*β* ± 1.96*se(HR_sd_)), where se(HR_sd_) is the standard error of the log-hazard ratio estimate. Risk factors have a multiplicative effect on the baseline hazard function. The model specifies the next τ-year absolute risk for a currently *a*-year old individual as

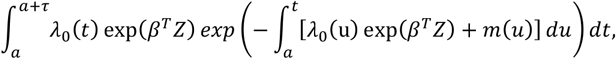

where *m(t)* is age-specific mortality rate function and λ_0_(*t*) is the baseline-hazard function, *t* ≥ *T* and *T* is the time to onset of the disease. The baseline-hazard function is derived from marginal age-specific BC incidence rates (*λ_m_*(*t*)) and distribution of risk factors *Z* in the general population (*F_z_*)).

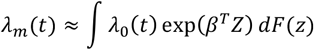

This absolute risk model allows disease background data from any country. In this analysis, we used Estonian background information. We calculated average cumulative risks using data from the National Institute of Health Development of Estonia (26) that provides population average disease rates in age groups of 5-year intervals. Sample sizes for each age group were acquired from Statistics Estonia for 2013-2016. Next, we assumed constant incidence rates for each year in the 5-year groups. Thus, incidence rates for each age group were calculated as *IR = Xt/Nt*, where Xt is the number of first-time cases at age *t* and N*t* is the total number of women in this age group. Final per-year incidences were averaged over time range 2013-2016.

Age-and sex-specific mortality data for the year 2016 was retrieved from World Health Organization (27) and competing mortality rates were constructed by subtracting yearly age- and sex-specific disease mortality rates from general mortality rates. Breast cancer mortality estimates were derived from the Global Cancer Observatory (1).

We applied this model to estimate absolute risks for individuals in the 1^st^, 10^th^, 25^th^, 50^th^, 75^th^, 90^th^ and 99^th^ PRS quantiles, eg. an individual on the 50^th^ percentile would have a standardized PRS of 0. Confidence intervals for the absolute risk are estimated with the upper and lower confidence intervals of the continuous per unit log-hazard ratio. Similarly, we used the absolute risk model to estimate lifetime risks (between ages 0 and 85) for the individuals in the same risk percentiles.

### PRS based risk-stratification and individual screening recommendations

Next, we evaluated PRS risk stratification in the Estonian BC screening context and simulated the extent of risk separation in the Estonian population. Females in Estonia currently start BC screening at age 50. Our analysis first established the 10-year risk of a 50-year old female with a population average of PRS (“average female”) using the model by Pal Choudhury (25): the reference for the level of risk that initiates population-level screening. Here, we assessed the differences in ages where individuals in various PRS risk percentiles attain 1 to 3-fold risk increases of risk compared to the 10-year risk of an average female.

Based on these analyses, we developed recommendations for a BC screening attendance program based on pre-screening PRS testing. This approach uses both relative risks, fold difference of 10-year risks compared to a genetically average woman of the same age, and also her absolute 10-year risk.

## Results

### Polygenic risk score re-validation in population cohort datasets

In the EGC cohort, we retained a total of 32,548 quality-controlled female samples. All samples were divided into prevalent and incident datasets. The prevalent dataset contained 315 cases of breast cancer that were diagnosed before Biobank recruitment and 1602 controls. The incident dataset contained 365 cases of breast cancer that were diagnosed after Biobank recruitment and 30,266 controls. The UKBB dataset contained 249,062 samples that passed the quality controls. In the UKBB, we identified 8637 prevalent cases and 6825 incident cases that were complemented with 44,952 controls and 188,648 controls, respectively.

Altogether, 4 models from 3 different articles were evaluated (9, 10, 28). Normality assumption of the standardized PRS was not violated with any tested models (Shapiro-Wilks test p-values in EGC data BC1 = 0.45, BC3 = 0.40, BC4 = 0.28, BC16 = 0.46). The best performing model was selected based on AUC, ORsd, AIC, and pseudo-R^2^ metrics in both EGC and UKBB data. The BC16 model that was based on Mavaddat et al. (9) performed the best (Table 1). The Corresponding AUC under the ROC curve (Figure 1) for the association between the PRS and BC diagnosis was 0.615 (SE = 0.039) in EGC and 0.632 (SE = 0.0072) in UKBB.

**Table 1.** Comparison metrics of BC PRS models based on the prevalent Estonian Genome Center dataset.

|  |  | BC1 (9) | BC3 (10) | BC4 (28) | BC16 (9) |
| --- | --- | --- | --- | --- | --- |
| Variants in original PRS |  | 313 | 77 | 78 | 3820 |
| Variants included in our model |  | 257 | 73 | 78 | 2803 |
| Estonian Genome Center | AUC (SE) | 0.604 (0.039) | 0.591 (0.004) | 0.573 (0.039) | 0.615 (0.039) |
| | $OR_{sd}$ ((SE [$\log OR_{sd}$])) | 1.43 (0.06) | 1.37 (0.06) | 1.27 (0.06) | 1.47 (0.06) |
|  | AIC | 1681.2 | 1689.6 | 1701.7 | 1674.1 |
|  | McFadden / Nagelkerke Pseudo-R<sup>2</sup> | 2.1% / 3.1% | 1.9% / 2.0% | 0.8% / 1.3% | 2.5% / 3.7% |
| UK Biobank | AUC (SE) | 0.625 (0.0073) | 0.607 (0.0075) | 0.584 (0.0073) | 0.632 (0.0072) |
|  | OR<sub>sd</sub> ((SE [log OR<sub>sd</sub>])) | 1.55 (0.012) | 1.48 (0.012) | 1.36 (0.012) | 1.62 (0.012) |
|  | AIC | 45 900 | 46 273 | 46 705 | 45 695 |
|  | McFadden / Nagelkerke Pseudo-R<sup>2</sup> | 3.0% / 4.5% | 2.2% / 3.3% | 1.3% / 2.0% | 3.5% / 5.1% |

**Figure 1.**
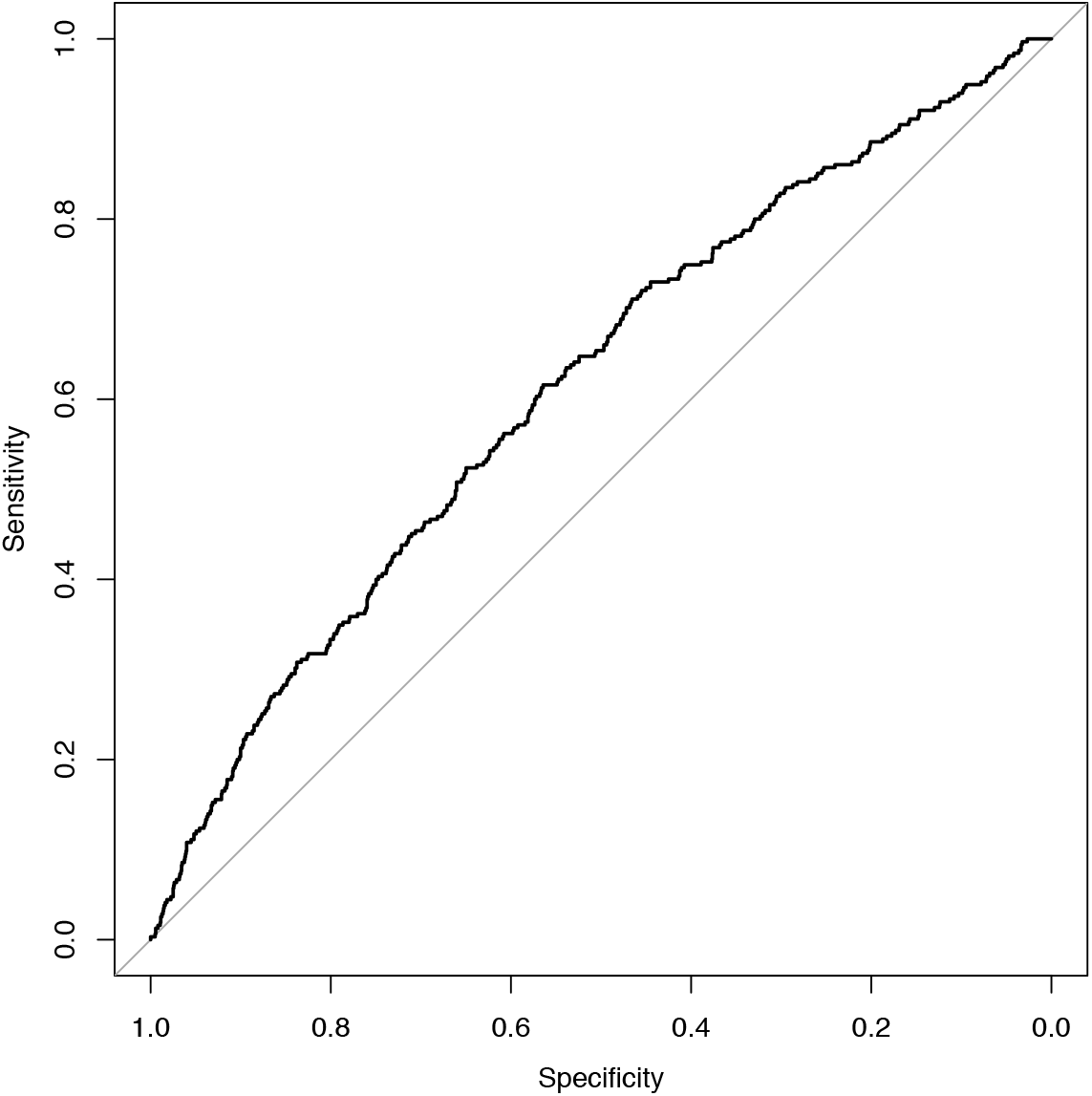
ROC plot of BC cases and controls in prevalent Estonian Genome Center dataset.

Next, we evaluated the performance of the best performing BC16 model in the independent incident datasets with the main aim of estimating the hazard ratio per unit of PRS. Table 2 presents the performance estimation metrics. Hazard ratio per 1 unit of standard deviation (HRsd) of model BC16 was *1.66* with standard error (log (*HR*)) = 0.05) in the incident EGC dataset. The concordance index (C-index) of the survival model testing the relationship between PRS and BC diagnosis status in the incident EGC dataset was 0.656 (se = 0.015) and slightly lower in the UK Biobank.

**Table 2.** Performance metrics of Cox regression model on the disease status and BC16 based polygenic risk scores calculated in the incident datasets.

|  |  | HR<sub>sd</sub> (95% confidence interval) | C-index (SE) | -2 × log likelihood | Likelihood ratio test p-value |
| --- | --- | --- | --- | --- | --- |
| BC16 (9) | Estonian Genome Center | 1.66 (1.5 – 1.84) | 0.656 (0.015) | 95.3 | < 2e-16 |
|  | UK Biobank | 1.56 (1.53-1.6) | 0.625 (0.003) | 1351 | < 2e-16 |

Hazard ratio estimates compared to individuals in the 40-60 percentile of PRS are visualized in Figure 2. In panel A with EGC data, the lowest 5% bin includes 1496 controls and 9 cases, whereas in the top 5%, we observed 1490 controls and 48 cases. With EGC the theoretical hazard ratio matched empirical estimate’s confidence intervals in 14 out of 16 comparisons. Alternatively, in panel B with UKBB data, the Spearman correlation coefficient between the empiric and theoretical hazard ratio estimates is 0.996, indicating very strong association, and the much narrower confidence bands match in 15 out of 16 cases.

**Figure 2.**
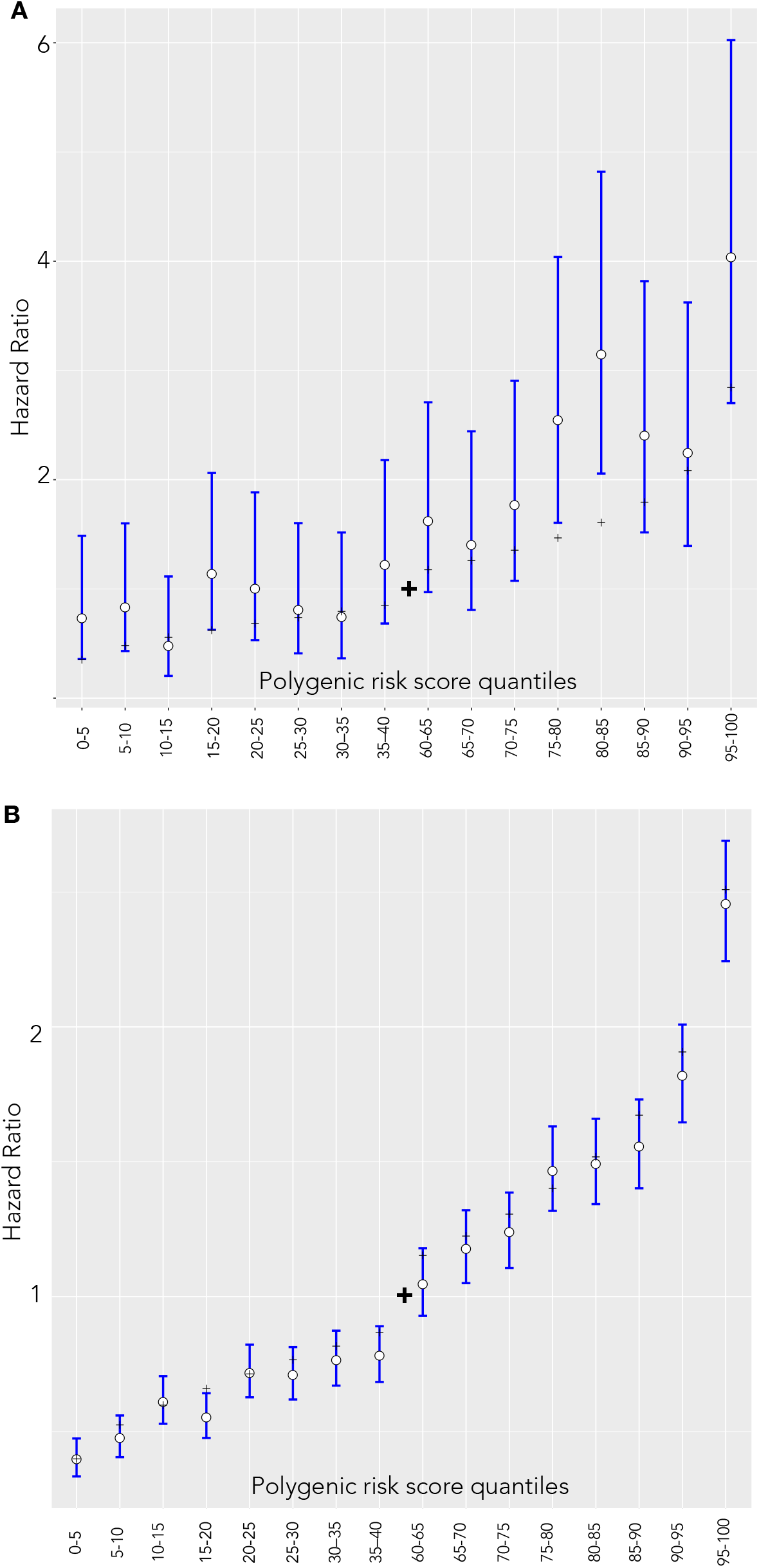
Hazard ratio estimates between quantiles 40–60 of the PRS and categorized 5% bins in the incident dataset. White dots and blue lines represent empirically estimated hazard ratio estimates and corresponding confidence intervals. Black dashes represent the theoretical hazard ratio for the 5%-quantile bins derived from the hazard ratio of per unit PRS. (A) Estonian Genome Center (B) UK Biobank.

### Polygenic risk score in breast cancer screening stratification

We used a model by Pal Choudhury et al. to derive individual 10-year risks (25) and specified *F(z)* as the distribution of PRS estimates in the whole EGC cohort. The log-hazard ratio (*β*) is based on the estimate of the log-hazard ratio in the BC16 model of the incident EGC dataset. Age-specific BC incidence and competing mortality rates provided the background for BC incidences in the Estonian population.

In the Estonian population, the absolute risk of developing breast cancer in the next 10 years among 50-year old women in the 1^st^ percentile is 0.466% (0.349% – 0.616%) and 4.83% in the 99^th^ percentile 4.83% (4.00 – 5.77%). At age 70, corresponding risks become 0.59% (0.445% – 0.778%) and 6.08% (5.03% – 7.30%) respectively. The relative risks between the most extreme percentiles are therefore > 10.3x fold. At the same time, competing risk accounted cumulative risks reach 19.2% by age 85 (16.1% – 22.6%) for those in the 99^th^ percentile but remain 2.00% (1.51% – 2.64%) for those in the 1^st^ percentile (Figure 3).

**Figure 3.**
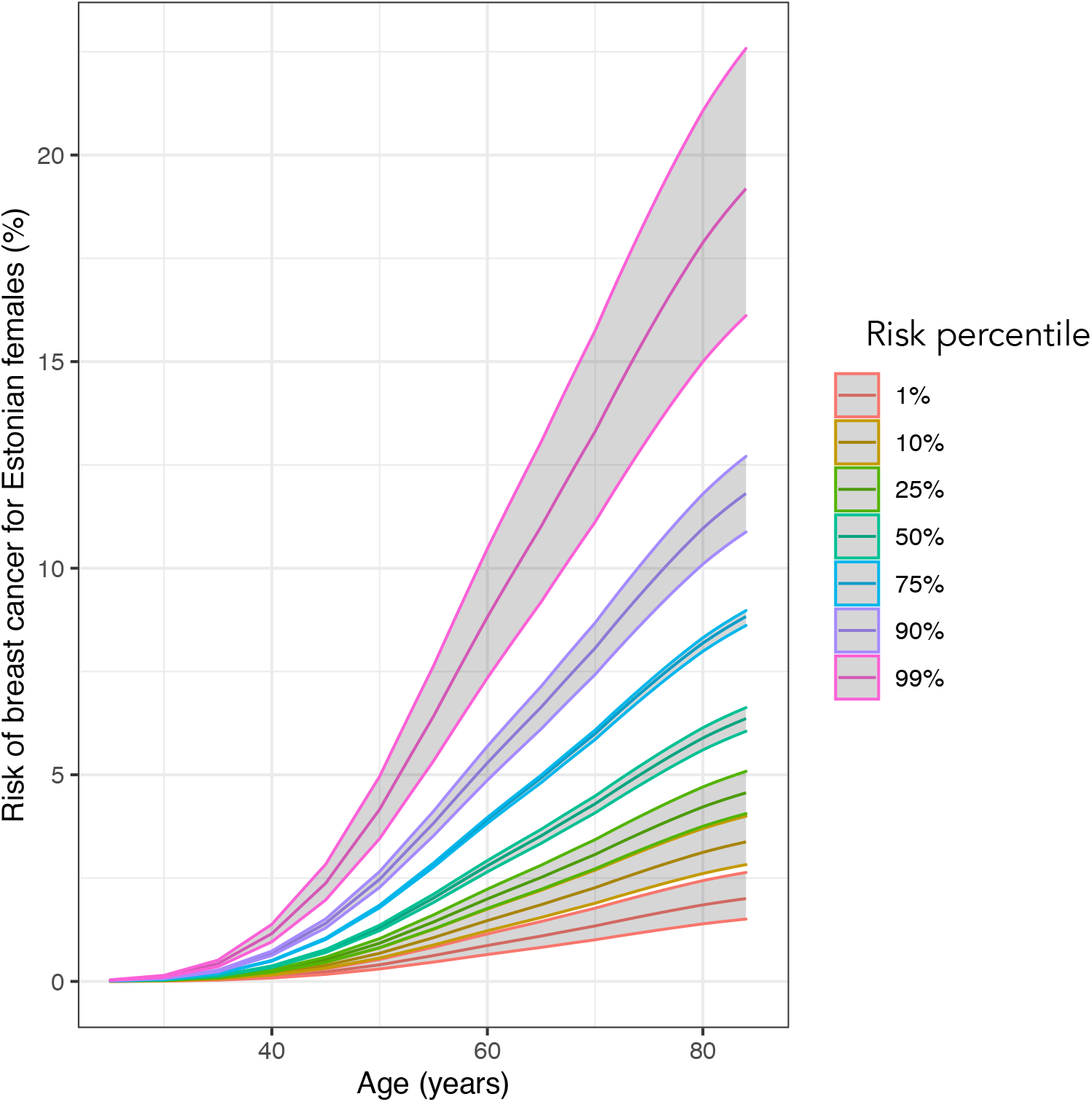
Cumulative risks (%) of BC between ages 20 to 85 in various risk percentiles.

A genetically average 50-year old female has a 10-year absolute risk of around 1.51%. BC16 model can identify 34-year-old females in the 99^th^ percentile of PRS that have a larger risk than this average risk of 50-year-olds. At the same time, 50-year-old females in the 32^nd^ percentile and lower attain average risk of 50-year-olds by their 70^th^ birthday. In effect, individual women could be at the risk that currently initiates population-level screening between ages 34 and 70. Similarly, 50-year old females above 92^nd^ PRS percentile have a more than 2-fold risk and around 1.3% of females attain a 3-fold risk compared to those at average risk (Figure 4).

**Figure 4.**
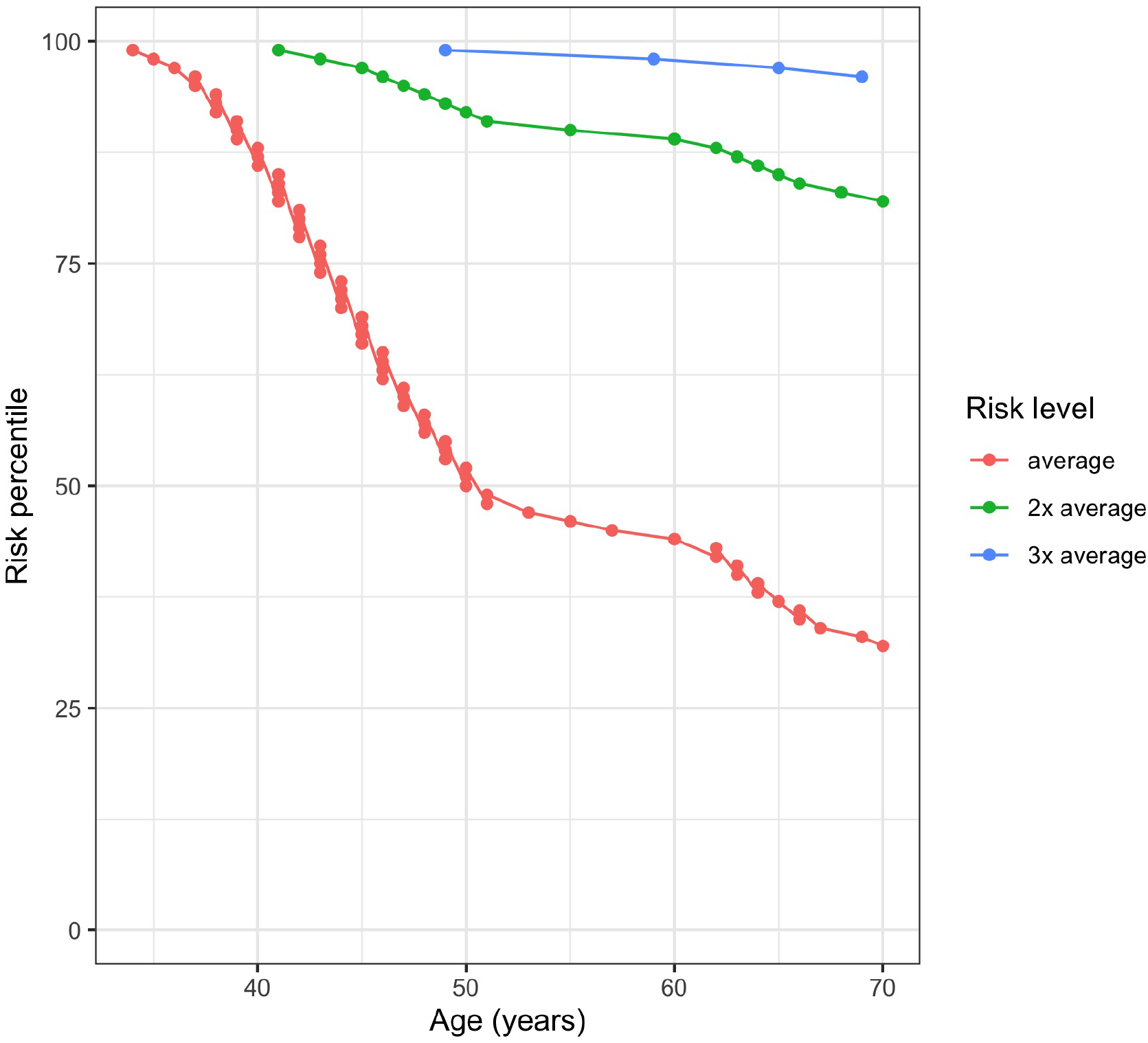
Ages when Estonian females in different risk percentiles attain 1-3 fold multiples of 10-year risk compared to 50-year old females with population average PRS (Risk level: “average”).

Based on PRS induced risk differences, we developed personalized recommendations that are based on relative risks compared to an individual of the same nationality, age, and sex, and also the estimated absolute risks. Recommendations presented below are based on the age when an individual attains the risk of genetically average 50-year old women. Mammography attendance procedures are also accompanied by general guidelines for reducing the risk of BC. It should be noted that we do not recommend individuals to join public screening programs later than standard starting time as the potential benefits and losses from decreased intervals have not been separately validated.

1. Relative risk is less than the population average

a. Standard mammography screening starting from age 50*
2. Relative risk increases up to 2-fold

a. Recommended mammography screening initiation (2-year interval) at the age of attaining 10-year risk equivalent to that of a genetically average 50-year old*
3. Relative risk increases 2 to 3-fold

a. Recommend 2a
b. Recommend mammography screening initiation with 1-year interval starting from an age where 10-year risk attains 2-fold of a genetically average 50-year old’s risk*
c. Besides to 3.a and 3.b, we recommend discussing the usage of BC risk-reducing hormonal chemoprevention (tamoxifen, aromatase inhibitors) with a specialist
4. Relative risk increases more than 3-fold

a. Recommend 3a and 3b
b. At the age of attaining more than 3-fold of a genetically average 50-year old’s risk then recommend magnetic resonance imaging (MRI) every 1-2 years*
c. Besides, to 4.a and 4.b, we recommend discussing the usage of BC risk-reducing hormonal chemoprevention (tamoxifen, aromatase inhibitors) with a specialist

* *If the recommended age is below the individual’s current age, then recommend current age*

## Discussion

Numerous full clinical prediction models incorporate traditional risk factors such as demographics, reproductive history, menopausal status, family history, previous biopsies, carrier status of high-risk monogenic mutations and mammographic density (29, 30). The practical routine application of such compounded models is complicated due to the non-availability and quality of data about individual risk factors. In practical settings, the data collection difficulties need to be weighed with expected gains. PRS alone has been shown to predict the risk of BC in European descent individuals more accurately than current clinical models (31). Additionally, Maas et al. looked at a large number of factors in risk stratification that yielded average absolute risk from 4.4% to 23.5% for women in the bottom and top 10% of risk distribution. ROC curves for prediction models showed a relatively modest increase of AUC for the model including a large number of clinical factors and PRS, compared to the PRS only model from 0.623 to 0.648 (20). Läll et al. demonstrated that the inclusion of family history improved the performance by around 1% compared to the PRS only model (32). Carriers of monogenic mutations such as BRCA1 have a severely increased lifetime risk but testing only provides utility for less than 1% of individuals. PRS is based on common variation and provides a single best estimate based on an unmodifiable measurable for all females, thus, incorporating PRS to screening strategy is reasonable.

In this study, we validated different publicly available PRS models to find the best performing model for predicting the risk of breast cancer. Models in this analysis have been previously validated in Estonia by Läll *et al*. (32). They compared several PRS risk scores including Mavaddat et al. (10) and Michailidou et al. (11) and their combinations in two population-based biobank cohorts. The best model by Läll et al. yielded a hazard ratio of 1.65 (95% CI 1.48 to 1.86) with a corresponding AUC of 0.636 (32). Mavaddat et al. (BC16, 3820 SNPs) estimated the mean of odds ratio for overall BC in several independent populations as 1.66 (95%CI: 1.61–1.79) and area under receiver-operator curve (AUC) = 0.636 (95%CI: 0.628–0.651 (9). In the UK Biobank, the estimated hazard ratio (HR) for overall BC per unit PRS for another model included in this study (BC1, 313 SNPs) was HRsd = 1.59. These risk discrimination results are consistent with our estimates.

Our best-performing model, named BC16, was a pruned version of Mavaddat et al 3820 PRS model containing a total of 2803 SNPs out of 3820. Its performance was consistent with the author’s results. Our model was used to design a novel absolute risk-based screening strategy. It is based on Estonian screening information and background data to identify the extent of more than 10-fold PRS-based risk differences between the extremes. Our analysis showed that one percent of women would need to join screening by the age of 34 and more than 30% of individuals do not ever attain the risks of a genetically average 50-year old woman (the age when women conventionally starting screening).

Our approach is easily adaptable to other nationalities by using population background information data of other genetically similar populations. Similarly, the clinical screening recommendations can be adapted to locality specific screening environments as long as we can infer the absolute risk of the average female in that locality.

In conclusion, we have used a PRS based model to develop a novel model for BC screening. Our adapted model identifies individuals at more than 3-fold risk and elucidated large differences in attaining the same level of absolute risk. The genetic risk-based recommendations can be applied prospectively by individuals and also by institutions aiming to make screening provision more efficient.

## Data Availability

Individual level genotype and phenotype data from Estonian Biobank or UK Biobank can not be explicitly shared. The UK Biobank Resource was used under Application Reference Number 53602. New users can request access to UK Biobank from http://www.ukbiobank.ac.uk/resources/.
Similarly, Estonian Biobank data is available by request. Researchers interested in Estonian Biobank can request the access here: https://www.geenivaramu.ee/en/access-biobank

https://www.geenivaramu.ee/en/access-biobank

## Acknowledgments

This research has been conducted using the UK Biobank Resource under Application Reference Number 53602 and with support by EIT Health The Digital Sandbox program.

Our appreciation goes to everybody from the Estonian Genome Center actively involved in this project. Specifically, we would like to thank Kristi Läll for recommendations on the manuscript and proofreading, prof. Krista Fischer for feedback in the study planning phase, Reidar Anderson, Merli Saare and Reedik Mägi for supporting the data exchange and running the analyses in EGC, and our friends Viljo Soo and Mari Nelis for great work in the laboratory. Also, we would like to thank prof. Lili Milani, prof. Andres Metspalu and prof. Tõnu Esko for their trust and support in this initiative.

Also, we would not have been able to perform any analyses without the computation resources provided by Sander Kuusemets and Ivar Koppel from the HPC Center of the University of Tartu.

## Ethical approval

### Estonian Genome Center

All human research was approved by the Research Ethics Committee of the University of Tartu, and conducted according to the Declaration of Helsinki and Human Research Act. All participants provided written informed consent to participate in the Estonian Biobank.

### UK Biobank

The UK Biobank study was approved by the North West Multi-Centre Research Ethics Committee (UK Biobank reference: 16/NW/0274). All participants provided written informed consent to participate in the UK Biobank study.

## Statement of data availability

Individual level genotype and phenotype data from Estonian Biobank or UK Biobank can not be explicitly shared. The UK Biobank Resource was used under Application Reference Number 53602. New users can request access to UK Biobank from http://www.ukbiobank.ac.uk/resources/. Similarly, Estonian Biobank data is available by request. Researchers interested in Estonian Biobank can request the access here: https://www.geenivaramu.ee/en/access-biobank

